# Comprehensive analysis of immune responses in CLL patients after heterologous COVID-19 vaccination

**DOI:** 10.1101/2022.09.21.22280205

**Authors:** Hye Kyung Lee, Manuela A. Hoechstetter, Maike Buchner, Trang Thu Pham, Jin Won Huh, Katharina Müller, Sabine Zange, Heiner von Buttlar, Philipp Girl, Roman Wölfel, Lisa Brandmeier, Lisa Pfeuffer, Priscilla A. Furth, Clemens-Martin Wendtner, Lothar Hennighausen

**Affiliations:** National Institute of Diabetes, Digestive and Kidney Diseases, National Institutes of Health, Bethesda, MD 20892, USA; Munich Clinic Schwabing, Academic Teaching Hospital, Ludwig-Maximilian University (LMU), Munich, Germany; Institute of Clinical Chemistry and Pathobiochemistry, School of Medicine, Technical University of Munich, Munich, Germany; TranslaTUM - Central Institute for Translational Cancer Research, Technische Universität München, 81675 Munich, Germany; Department of Pulmonary and Critical Care Medicine, Asan Medical Center, University of Ulsan College of Medicine, Seoul, Korea; Bundeswehr Institute of Microbiology, Munich, Germany; German Centre for Infection Research (DZIF), Partner Site Munich, Munich, Germany; Departments of Oncology & Medicine, Georgetown University, Washington, DC, USA

## Abstract

Patients with chronic lymphocytic leukemia (CLL) treated with B-cell pathway inhibitors and anti-CD20 antibodies exhibit low humoral response rate (RR) following SARS-CoV-2 vaccination. To investigate the relationship between the initial transcriptional response to vaccination with ensuing B and T cell immune responses, we performed a comprehensive immune transcriptome analysis flanked by antibody and T cell assays in peripheral blood prospectively collected from 15 CLL/SLL patients vaccinated with heterologous BNT162b2/ChAdOx1 with follow up at a single institution. The two-dose antibody RR was 40% increasing to 53% after booster. Patients on BTKi, venetoclax ± anti-CD20 antibody within 12 months of vaccination responded less well than those under BTKi alone. The two-dose T cell RR was 80% increasing to 93% after booster. Transcriptome studies revealed that seven patients showed interferon-mediated signaling activation within 2 days and one at 7 days after vaccination. Increasing counts of COVID-19 specific IGHV genes correlated with B-cell reconstitution and improved humoral RR. T cell responses in CLL patients appeared after vaccination regardless of treatment status. A higher humoral RR was associated with BTKi treatment and B-cell reconstitution. Boosting was particularly effective when intrinsic immune status was improved by CLL-treatment.

## Introduction

Patients with chronic lymphocytic leukemia (CLL) are considered high-risk for severe COVID-19 infection, mainly due to their complex underlying immunodeficiency and inadequate immune response to infections.^1-3^ They not only suffer from immune dysregulation by the disease itself, but their immune system is further disrupted by treatment-related effects.^4-6^ Patients, who are heavily pre-treated with chemoimmunotherapy, exposed to anti-CD20 antibody or treated actively with B-cell pathway inhibitors, experience suboptimal antibody response to COVID-19 vaccination compared to CLL treatment-naïve.^7-13^ Robust data on immunogenicity of 2-dose homologous or heterologous BNT162b2/ChAdOx1 vaccine schedules in leukemia patients have demonstrated an enhanced humoral and/or cellular immune response.^14,15^ Heterologous vaccine schedules also enhance humoral response in individuals without hematological disease.^16,17^ The European Medicines Agency (EMA) and the European Centre for Disease Prevention and Control (ECDC) discussed potential benefits of heterologous regimens in 2021.^18^

While CLL patients who received their last treatment within 12 months preceding standard vaccination program demonstrate low response rates, vaccine response rates increase in seronegative, actively treated patients following boosting.^14,19^ In addition, potential protection against COVID-19 infection provided by T cells, even in the absence of a humoral response, is of particular clinical interest.^20,21^ T cell activation with release of IFN-γ by SARS-CoV-2 is associated with mild disease and viral clearance.^21,22^ T cell mediated immune responses are reported in patients with lymphoid malignancies in the absence of a humoral response. ^23^ However, in a mixed group of cancer patients, they were documented more commonly in combination with a humoral response.^24^

Early responses to vaccination are elevated levels of interferons and other cytokines, which activate the JAK/STAT signaling pathway and induce expression of immediate and innate response genes. We have used RNA-seq of peripheral immune cells to identify the innate immune response of healthy individuals receiving the standard homologous BNT162b2^25^ or a heterologous ChAdOx1/BNT162b2^17^ regimen. Specific genetic pathways are differentially activated within the first two days after vaccination and more prominently in the heterologous cohort. However, there are no reports in the literature on the immune transcriptomic response in CLL patients receiving COVID-19 vaccines.

The utility of heterologous vaccination regimens for improving immune response in immune compromised patients continues to be deliberated.^26,27^ To add to this important discussion, here we provide a comprehensive transcriptome analysis of peripheral immune cells from CLL patients who received heterologous ChAdOx1/BNT162b2 vaccination and monitored their innate and humoral immune response until four months following third vaccination in combination with detailed discussion of disease-status, treatment regimens, and response to COVID-19 infection during follow-up.

## Methods

### Ethical approval

Ethical approval (#20-225) to conduct this analysis was granted by the institutional review board of the Ludwig-Maximilian University (LMU), Munich as the responsible ethics committee. Written informed consent was obtained from the study participants.

### Study population, study design and recruitment

From June 2021 through July 2021, 15 patients diagnosed with CLL/SLL between 2003 and 2021 were recruited in a single institution (Department of Hematology and Infectious Diseases, Munich Clinic, Munich Schwabing, Germany), per recommendation of booster vaccination by the Standing Committee on Vaccination (STIKO) at the Robert Koch Institute in Germany. After providing written informed consent for data collection, five seronegative patients received a third (3-dose) after standard 2-dose homologous vaccination of BNT162b2 or ChAdOx1. All were heavily pretreated (103, 104) or recently treated with anti-CD20 mAbs (105, 106) or a BTK inhibitor (107). In addition, ten patients (2-dose) with different CLL disease and treatment status (201 to 213), half of whom were seropositive after prime dose of ChAdOx1, received a second homologous or heterologous dose. At the time of vaccination 14 of 15 patients did not have a history of COVID-19 infection. Between October 2021 and December 2021, all ten patients of 2-dose received a third BNT162b2 dose. Antibody response and incidence and outcome of COVID-19 infections were recorded per routine CLL management. Four patients had a breakthrough COVID-19 infection with Omicron variant around six months after the third vaccination, all with mild symptoms. One patient received antiviral treatment with molnupiravir. Patient #209 had an undiagnosed infection in January 2020, confirmed by anti-SARS-CoV-2 nucleocapsid IgG antibodies in follow-up.

### Stimulation of lymphocytes and detection of IFN-γ

For lymphocyte stimulation studies, heparinized blood samples were transported within four hours of collection to the laboratory. 1mL sample was then transferred to three QuantiFERON (QFN) ® SARS-CoV-2 (Qiagen) SARS-CoV-2 blood collection tubes (Sars-CoV-2 specific antigens AGI, AGII, AGIII). After 24 hours of stimulation, plasma from the stimulated samples was used for the detection Interferon gamma (IFN-γ). Detection was carried using the QuantiFERON ® ELISA Human IFN-γ (Qiagen). Detected IFN-γ level >0.1 IU/ml is evaluated as positive response.

### SARS-CoV-2 antibody ELISA

End-point binding IgG levels to the S1 domain of the spike protein of SARS-CoV-2 were measured using the semi-quantitative Anti-SARS-CoV-2 ELISA IgG (EUROIMMUN, Lübeck), according to the manufacturer’s instructions. Positive responses included both IgG ratio ≥ 1.1 and borderline values IgG ≥ 0.8 to <1.0. Negative responses were IgG ratio < 0.8. In addition, Quantitative Anti-SARS-CoV-2 ELISA IgG measurement (Atellica IM SARS-CoV-2 IgG, Siemens) was performed with positive response >= 21.8 Binding Antibody Unit/ml (BAU/ml) and negative < 21.8 BAU/ml.

### Virus Neutralization Test

SARS-CoV-2 (strain MUC IMB-1, clade B1) neutralizing antibody titers were determined as previously described^28^, including positive and negative controls. Heat-inactivated serum samples in duplicates, including positive and negative control samples, were serially diluted in 96-well tissue culture plates starting at 1:5 to a maximum of 1:640. Virus stocks (50 TCID/50 μl) were prepared and stored at −80 °C until further use. Virus was pre-incubated (1 h, 37 °C) with diluted serum samples before Vero E6 cells (1×10^4^ cells/50 μl) were added. After 72 h (37 °C), supernatants were discarded and wells were fixed (13% formalin/PBS) and stained (crystal violet, 0.1%). The neutralizing antibody titer corresponded to the reciprocal of the highest serum dilution showing complete inhibition cytopathic effect (CPE).

### Flow Cytometry

Cells were stained with fluorochrome-labeled antibodies to manufacturer information (Staining 1: CD19 PE-Cy7 and CD5 APC; Staining 2: CD3 FITC, CD4 APC-Cy7, CD8 Amcyan, PD1 PE-Cy7, CD25 PE, CD62L APC; staining 3: CD11c PE, CD14 APC-Cy7, HLA-DR PE-Cy7, CD56 PerCP710, all Biolegend). To block free Fc receptors human Fc Receptor Binding Inhibitor Polyclonal Antibody (eBioscience) were added 10 min prior to labeled antibodies. Dead cells were excluded by DAPI (1 µg/ml) (Sigma Aldrich) staining. Flow cytometric analysis was performed using a FACS Canto II cytometer (BD Bioscience). Data was analyzed with the FlowJo™ software version 10.7.1 (BD Bioscience).

### Extraction of the buffy coat and purification of RNA

Peripheral blood mononuclear cells (PBMCs) were isolated from whole blood by density gradient centrifugation using Ficoll-Paque (GE Healthcare, Chicago, IL, USA). CD19^+^ B-cells were depleted by magnetic-activated cell sorting (MACS) using human CD19 MicroBeads (Miltenyi, Bergisch-Gladbach, Germany) if CLL cell population exceeded 10% of viable lymphocytes, as determined by flow cytometry, prior to RNA extraction. 3×10e6 PBMC (with or without CD19 depletion) were collected, washed with PBS and resuspended in 200ul Homo-TG buffer (Maxwell® 16 LEV simplyRNA Purification Kit, Promega) and stored at −80°C. RNA was extracted on the Maxwell® 16 Instrument according to manufacturer’s protocol and stored at −80°C for further processing.

### mRNA sequencing (mRNA-seq) and data analysis

Bulk RNA-seq was performed on three million PBMCs obtained prior to the second vaccination (2-dose cohort) and third vaccination (3-dose cohort) and at days 1/2 (D1/2), 7 (D7) and week 4-5 (W4-5) after the vaccination. RNA-seq was conducted on a total of XX samples with an average sequencing depth of at least 200 million reads per sample. The Poly-A containing mRNA was purified by poly-T oligo hybridization from 1 mg of total RNA and cDNA was synthesized using SuperScript III (Invitrogen). Libraries for sequencing were prepared according to the manufacturer’s instructions with TruSeq Stranded mRNA Library Prep Kit (Illumina, RS-20020595) and paired-end sequencing was done with a NovaSeq 6000 instrument (Illumina) yielding 200-350 million reads per sample. The raw data were subjected to QC analyses using the FastQC tool (version 0.11.9) (https://www.bioinformatics.babraham.ac.uk/projects/fastqc/). mRNA-seq read quality control was done using Trimmomatic^29^ (version 0.36) and STAR RNA-seq^30^ (version STAR 2.5.4a) using 150 bp paired-end mode was used to align the reads (hg19). HTSeq^31^ (version 0.9.1) was to retrieve the raw counts and subsequently, Bioconductor package DESeq2^32^ in R (https://www.R-project.org/) was used to normalize the counts across samples and perform differential expression gene analysis. Additionally, the RUVSeq^33^ package was applied to remove confounding factors. The data were pre-filtered keeping only genes with at least ten reads in total. The visualization was done using dplyr (https://CRAN.R-project.org/package=dplyr) and ggplot2.^34^ The genes with log2 fold change >1 or <-1 and adjusted p-value (pAdj) <0.05 corrected for multiple testing using the Benjamini-Hochberg method were considered significant and then conducted gene enrichment analysis (GSEA, https://www.gsea-msigdb.org/gsea/msigdb). For T- or B-cell receptor repertoire sequencing analysis, trimmed fastq files from bulk RNA-seq were aligned against human V, D and J gene sequences using the default settings with MiXCR.^35,36^ CDR3 sequence and the rearranged BCR/TCR genes were identified.

### Statistical analysis

Differential expression gene (DEG) identification used Bioconductor package DESeq2 in R. P-values were calculated using a paired, two-side Wilcoxon test and adjusted p-value (pAdj) corrected using the Benjamini–Hochberg method. Genes with log2 fold change >1 or <-1, pAdj <0.05 and without 0 value from all sample were considered significant. For significance of each GSEA category, significantly regulated gene sets were evaluated with the Kolmogorov-Smirnov statistic. A value of \**p* < 0.05, \*\**p* < 0.01, \*\*\**p* < 0.001, \*\*\*\**p* < 0.0001 was considered statistically significant.

### Data Sharing Statement

The RNA-seq data from this study were deposited under the accession GSE201642 in the Gene Expression Omnibus (GEO). RNA-seq data of healthy heterologous vaccinated individuals were obtained under GSE201535.

## Results

### Patient characteristics

Patient characteristics are shown in Table 1, Figure 1A and Supplementary Table 1. At the time of the vaccination, two patients (13%) had treatment-naïve CLL. Seven (47%) were on treatment without remission (2 frontline, 4 relapse) or with remission (1 relapse). Six (40%) were off therapy, including four in clinical complete or partial remission (2 frontline, 2 relapse) and two on relapse in need of treatment (1 frontline, 1 relapse). Of the treated patients, four received venetoclax monotherapy and three ibrutinib monotherapy. Eleven of 15 patients were previously on anti-CD20 monoclonal antibodies with/without chemotherapy, either more than 12 months (7 patients) or within 12 months (4 patients) prior to vaccination. Unfavorable prognostic CLL parameters included β2M >3.5 mg/l (2/15), complex karyotype (2/15), trisomy 12 (1/15), unmutated *IGHV* gene status (13/15), presence of TP53/del(17p) and/or del(11q) (6/15). Median IgG level was 619 mg/dl (range 159-1141), IgM level of 45 mg/dl (range <5-179) and IgA level of 98 mg/dl (range 12-210). Median ALC (absolute lymphocyte count) was 5.9/μl (range 3.3-46.7).

**Table 1.**
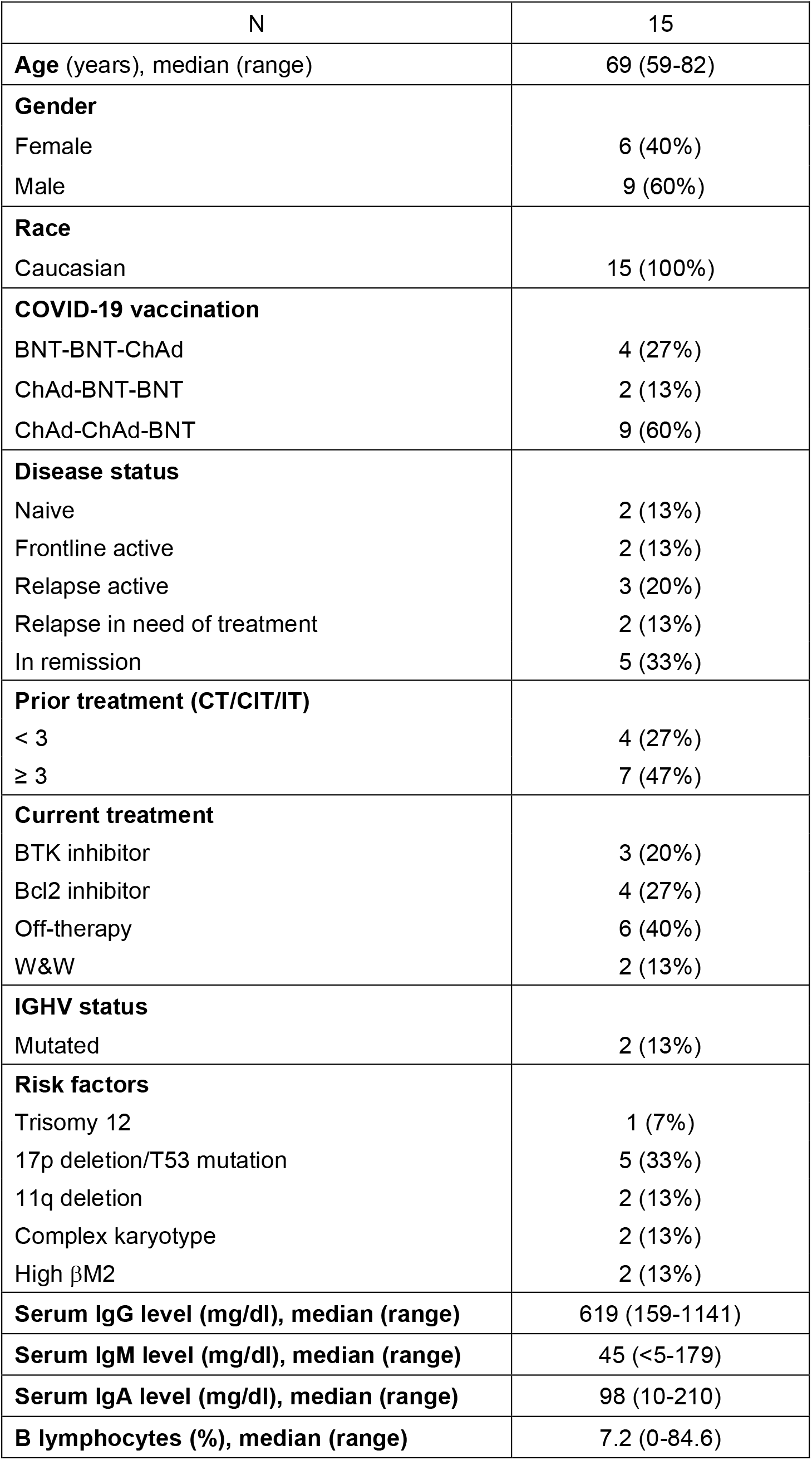
Characteristics of CLL study population.

**Figure 1.**
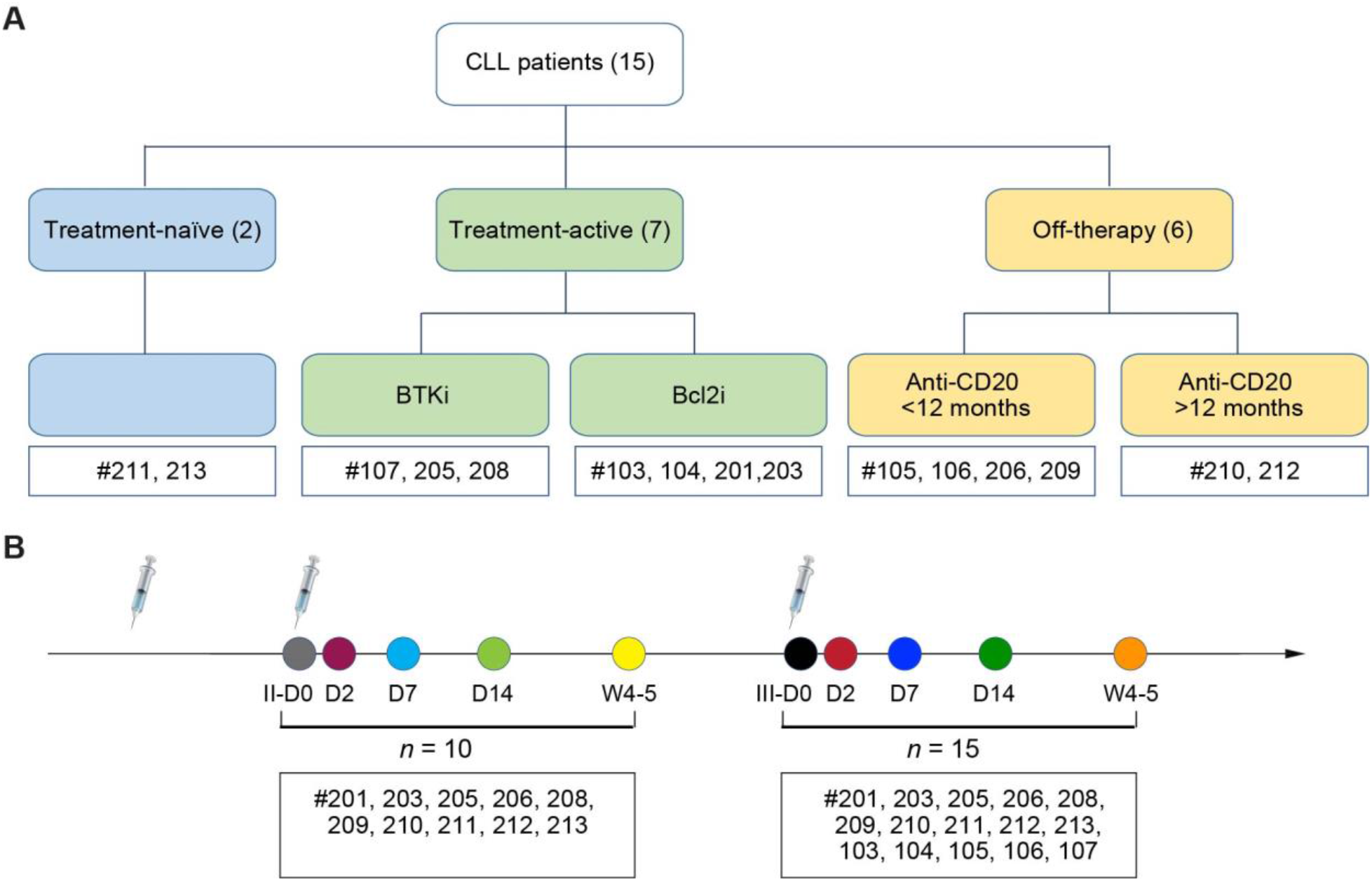
Study population and design. (A) CLL patient groups by treatment (naïve (n=2), treatment active (n=7), off-treatment (n=6)) and by drugs (BTKi (n=3), Bcl2i (n=4), anti-CD20 for less than 12 months (n=4), anti-CD20 for more than 12 months (n=2)). Mean distances between dose 1 and 2, and dose 2 and 3 were 82 and 132 days, respectively. (B) Blood samples were collected prior to the vaccination (D0) and at days 2 (D2), 7 (D7) and 14 (D14) and weeks 4-5 (W4-5) post second and third vaccination as indicated by the colored circles.

### Antibody response

We analyzed anti-spike IgG antibody levels, neutralizing antibody levels and immune transcriptomes (RNA-seq) on peripheral immune cells prior to the vaccination (referred to as Day (D) 0 (D0), on days 2 (D2), 7 (D7) and 14 (D14) and weeks 4-5 (W4-5) post vaccination (Figure 1B). Circulating antibody levels were most closely correlated with treatment history.^9,37^ After 2-dose vaccination, anti-spike (S) IgG was detected in 6/15 (40%) patients, including 2/6 (33%) off-therapy (#210, 212); 2/2 (100%) treatment-naïve (#211, 213); 1/4 (25%) on active venetoclax (#203); 0/3 (0%) on ibrutinib; 1/4 (25%) on anti-CD20 of less than 12 months (#209) (Figures. 2A and 3A; Supplementary Table 2). Following three vaccination doses, 8/15 patients (53%) showed detectable anti-S antibodies (Figures 2A and 3A; Supplementary Table 2). Several interesting responses occurred after a three-dose heterologous regimen. Two naïve patients (#203, #209) that received frontline therapy with acalabrutinib shortly before or after their third BNT162b2 dose retained seroconversion beyond 6 months post treatment. Patient #203 received frontline treatment with venetoclax monotherapy prior to first ChAdOx1 vaccination, showed a detectable anti-S IgG response at D14 post second ChAdOx1 dose that was lost at W4-5 but then restored after a third BNT162b2 dose. Patient #209, who had an undiagnosed COVID-19 infection prior to receiving first vaccination and stopped venetoclax+anti-CD20 treatment shortly before the second vaccination, showed an antibody response after first vaccination and retained detectable antibody levels through subsequential vaccination even with ongoing B-cell depletion in the context of CLL remission. It is reported that SARS-CoV-2 infection leads to more robust and durable B and T cell immune responses than COVID-19 vaccination in CLL patients.^38^ Two patients on ibrutinib (#107, 205), who failed humoral response after two homologous ChAdOx1 doses, seroconverted after the third heterologous BNT162b2 dose. Patient #107, who was in second relapse and on ibrutinib treatment for two years, showed a delayed seroconversion three months after a temporal pause of ibrutinib, while receiving a third heterologous BNT162b2 dose. Patient #205, who was on ibrutinib within 6 months, showed a delayed antibody response to the third heterologous BNT162b2 vaccination. Results were consistent with the notion that patients with adequate levels of serum immunoglobulins showed an increased antibody response after third vaccination, whereas those with very low serum levels of IgA, IgG and IgM, respectively, failed to seroconvert following vaccination (Supplementary Table 1). Overall, 7/15 of patients failed to mount a detectable humoral response even after 3-dose vaccination, irrespective of homologous or heterologous vaccination protocol. These patients were all heavily pre-treated. Three were on current venetoclax treatment (#103, 104, 201), one presented with CLL relapse with therapy pending (#106), one on prolonged ibrutinib (#208), and two with anti-CD20 exposure less than 12 months before vaccination (#105, 206). Development of neutralizing antibodies was limited to the four patients showing a maximal antibody response following 2-dose vaccination that was not boosted by 3-dose vaccination (# 210, 211, 212, 213) (Figure 2B; Supplementary Table 2).

**Figure 2.**
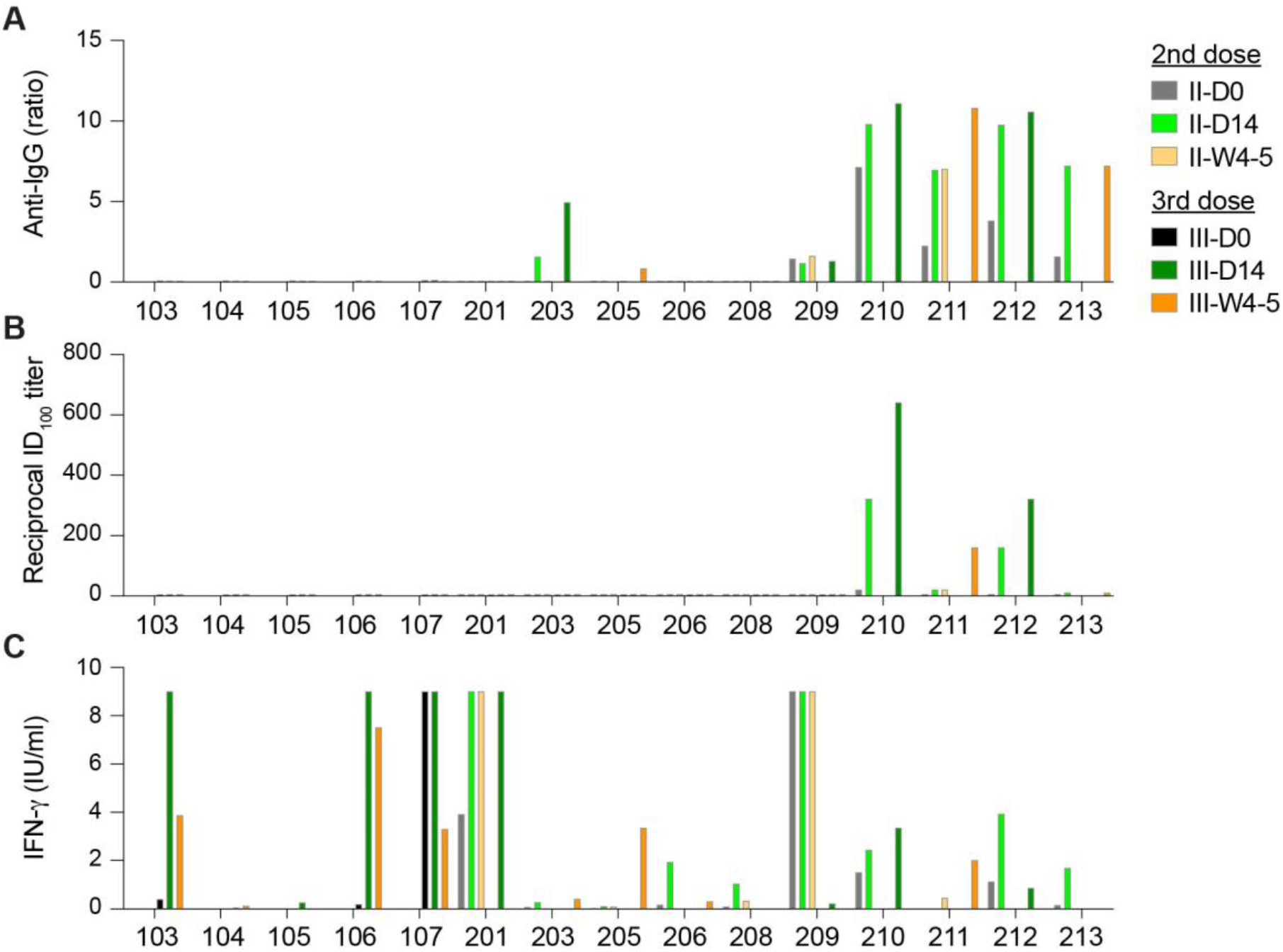
Humoral and T cell immune responses following second dose and booster vaccination. (A) Plasma IgG antibody binding against the S1 domain of SARS-CoV-2 spike in CLL patients. (B) Neutralizing antibody response to SARS-CoV-2 RBD (spike). (C) IFN-γ levels secreted from lymphocytes by stimulation of a SARS-CoV-2 peptide cocktail.

### Cellular immune response

Robust T cell responses were detected in 12/15 patients (80%) after 2-dose vaccination and 14/15 patients (90%) following 3-dose vaccination (Figure 2C; Supplementary Table 3). T-cell response rate was independent of clinical characteristics, intrinsic immune and treatment status (Figure 3B). All seropositive patients (8/8) as well as 6/7 seronegative patients developed a T cell mediated IFN-γ response. The one seronegative patient (#104) without a T cell response even after a third heterologous vaccination was on current venetoclax treatment. One patient (#213), who started frontline therapy with acalabrutinib shortly before the third heterologous vaccination, lost T cell response. Further analysis of cellular immune cell activation in all patients via flow cytometry revealed highly variable levels of activated and exhausted T cells, myeloid derived-suppressor cells (MDSCs), and NK cells was performed but there was no correlation between these results and response rates detected by IFN-γ (Supplementary Table 4).

**Figure 3.**
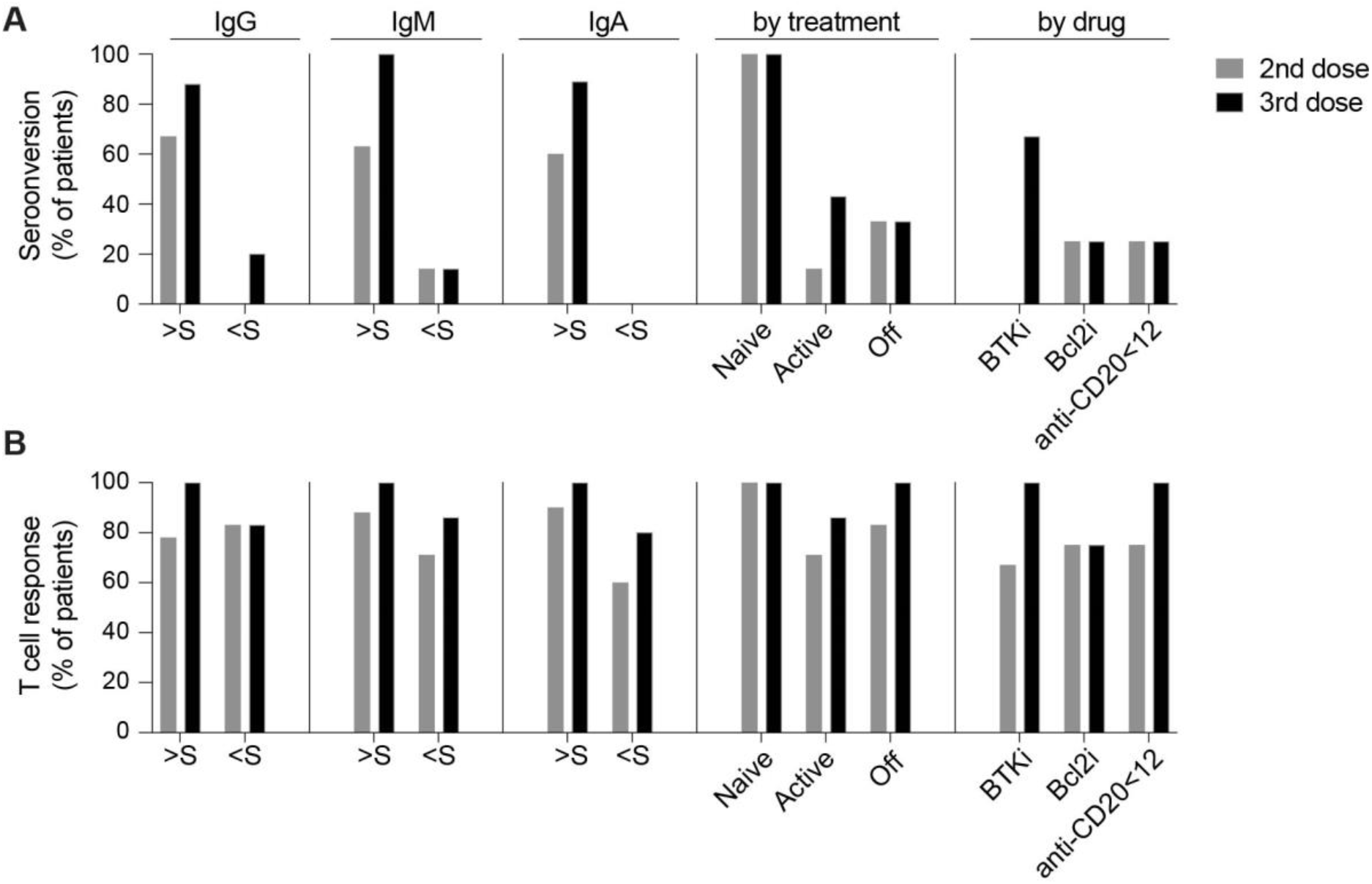
Anti-spike mediated T cell response in CLL patients. (A) Seroconversion rate. (B) T cell response rate after 2-dose and 3-dose vaccines in patient subgroups based on clinical parameters and treatment status. Y-axis presents the proportions (%) of CLL patients. S, standard of active level, IgG: 552 mg/dl, IgM: 33 mg/dl, IgA: 69 mg/dl.

### Transcriptional immune response

Next, we assessed the vaccine-induced innate immune response and transcriptional response in PBMCs isolated from 2- and 3-dose vaccination cohorts at days 0 (D0), 2 (D2), 7 (D7) and week 4-5 (W4-5) following vaccination (Figure 1B). The sequencing depth of 200 million reads per sample permitted an in-depth analysis of early response immunes and germline alleles induced by the vaccine. Reference cohorts were healthy individuals receiving heterologous or homologous vaccinations.^17^ Samples were available for eight patients: 3-dose BNT-BNT-ChAd (#103, #104, #106), 2-dose ChAd-ChAd (#201, #208, #209), 2-dose ChAd-BNT (#206, #212). First, numbers of differentially expressed genes (DEGs) were measured to examine the immediate response upon vaccination (Figure 4; Supplementary Figures 1-2). IFN-γ enrichment scores for DEGs at D2 were independent of antibody response. High levels were found both in patients with (#209, #212) and without (#201, #208) an antibody response (Figure 4A). Cell cycle pathway enrichment scores for DEGs were highest at D7, significantly higher both as compared to D2 (Figure 4B) and D0 (Figure 4C). Similarly, higher enrichment in cell cycle pathways also did not correlate with seroconversion (#106, #201, #208). Patient #104, the one patient without a T-cell response, showed relatively low enrichment for both IFN-γ enrichment scores at D2 and cell cycle pathway enrichment at D7. Patient #206 had a higher IFN-γ score at D7 compared to D2, suggesting delayed immune response. This patient exhibited a T-cell response and experienced a mild COVID-19 infection during follow-up, but this was not associated with seroconversion. While the magnitude of activation varied, activation of interferon-induced genes (*IFI27, ISG15, CXCL10, GBP1*), JAK/STAT signaling genes including *STAT1*, antiviral pattern recognition receptors (*DDX58, DHX58*) and *OAS* family genes at D2 were qualitatively similar to healthy controls (Fig. 4D). Induction of *IFI6* and *IFIT* genes were higher in CLL patients compared to healthy controls. While the transcriptome response varied greatly between individual CLL patients and was independent of the clinical characteristics and treatment status, the vast majority of CLL patients regardless of antibody response exhibited an early transcriptome immune response presaging a later sustained T-cell mediated IFN-γ immune response.

**Figure 4.**
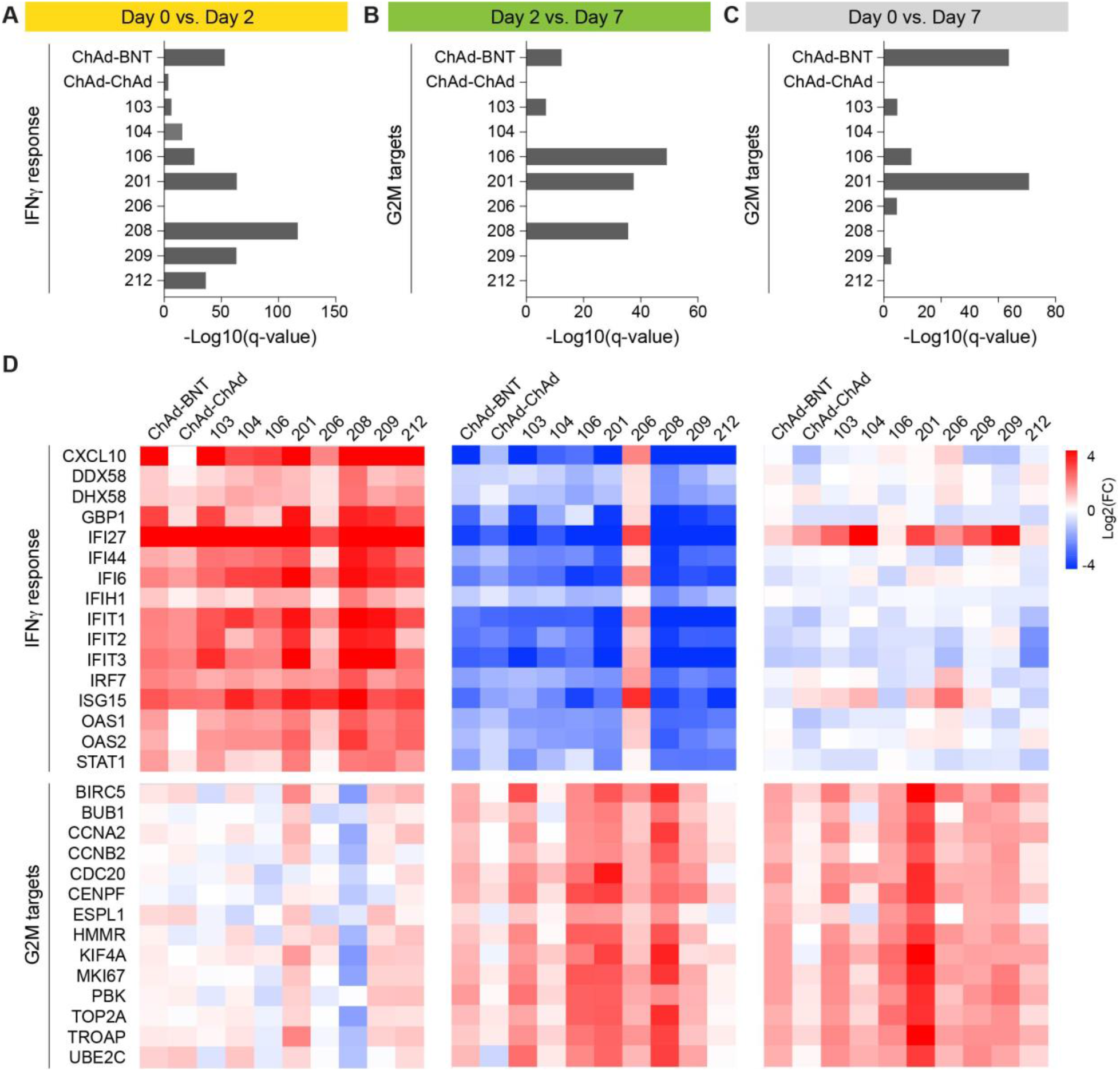
Immune transcriptomes following vaccination. (A-C) Interferon-regulated genes are induced upon vaccination. Genes expressed at significantly higher levels at days 2 and 7, as compared to day 0 were significantly enriched in Hallmark gene sets. X-axis denotes statistical significance as measured by minus logarithm of FDR q-values. Y-axis denotes ranked terms by q-values. (D) Heatmaps show log2 FC of IFN-γ response genes (top) and G2M target genes (bottom) significantly regulated between day0 and day2, day2 and day7, and day0 and day7.

### Immunoglobulin germline repertoire

Finally, expression profiles of specific germline variable gene classes were interrogated. First, the range of immunoglobulin heavy chain variable (*IGHV*), light chain (*IGKV, IGLV*), and T cell receptor alpha/beta variable (*TRAV, TRBV*) gene usage in the CLL patients was assessed (Figures 5-6). Final clonotype numbers were more than four-fold lower at D0 and D7 as compared to vaccinated healthy controls (Figure 5A). A broad range of germlines in each patient was revealed by a deeper analysis of *IGHV* using complementarity determining regions (CDR)*1* and CDR2 (Figure 5B). We observed *IGHV3-74, IGHV3-30/IGHV3-33, IGHV1-18, IGHV3-23, IGHV3-21* and *IGHV4-59* that are the basis of neutralizing antibodies identified in SARS-CoV-2 patients.^39-42^ In six patients these clones were specifically increased (#103, #104, #201, #203, #212, #213). Three antibody responsive patients, including treatment-naïve (#213), on venetoclax (#203), and off-therapy (#212), showed relatively higher numbers of IGHV clones. Because of transiently diminished B-cells upon active venetoclax treatment, patient #203 had more IGHV clones at D0 and D7 than W4-5 while other antibody responsive patients (#209, #210, #211) showed low and progressive numbers by recovered B cells. Notably, no B cell receptor (*BCR*) clones were detected in the antibody non-responder #206, likely due to completely depleted B cells by CLL treatment. Overall, results illustrate initially low levels of *BCR* in CLL patients lacking a humoral and neutralizing antibody response gradually increase upon B-cell reconstitution with effective CLL therapy. In contrast to *BCR* genes, activation of T Cell Receptor (*TCR*) genes (*TRAV, TRBV*) was readily detected (Figure 6A), consistent with preservation of T cell responses. Several *TRAV* and *TRBV* genes that are present in COVID-19 convalescent patients^43,44^ were induced in the CLL patients at between D0 and W4-5 (Figure 6B).

**Figure 5.**
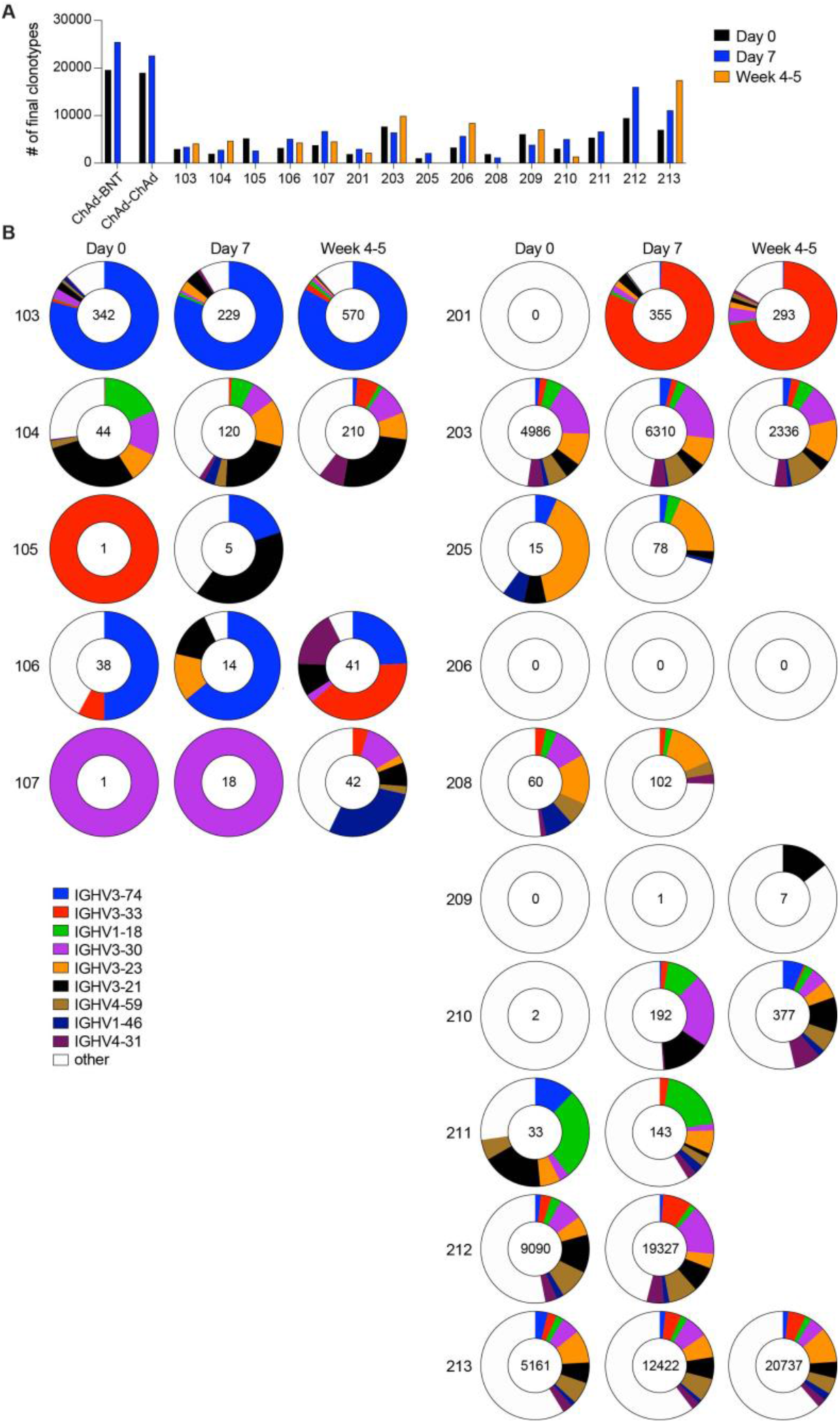
SARS-CoV-2 B-cell memory. (A) Bar graphs were shown the number of clonotypes in each patient after the vaccination. (B) Pie charts for IGHV show the distribution of antibody sequences of individuals prior to the vaccination and after 7 days and 4~5 weeks. The number of sequences analyzed for individual are shown in the inner circle. Sizes of pie slices are proportional to the number of clonally related sequences. Persisting clones (same IGV genes) in both time points are shown as colored slices. White indicates sequences not overlapped between individuals.

**Figure 6.**
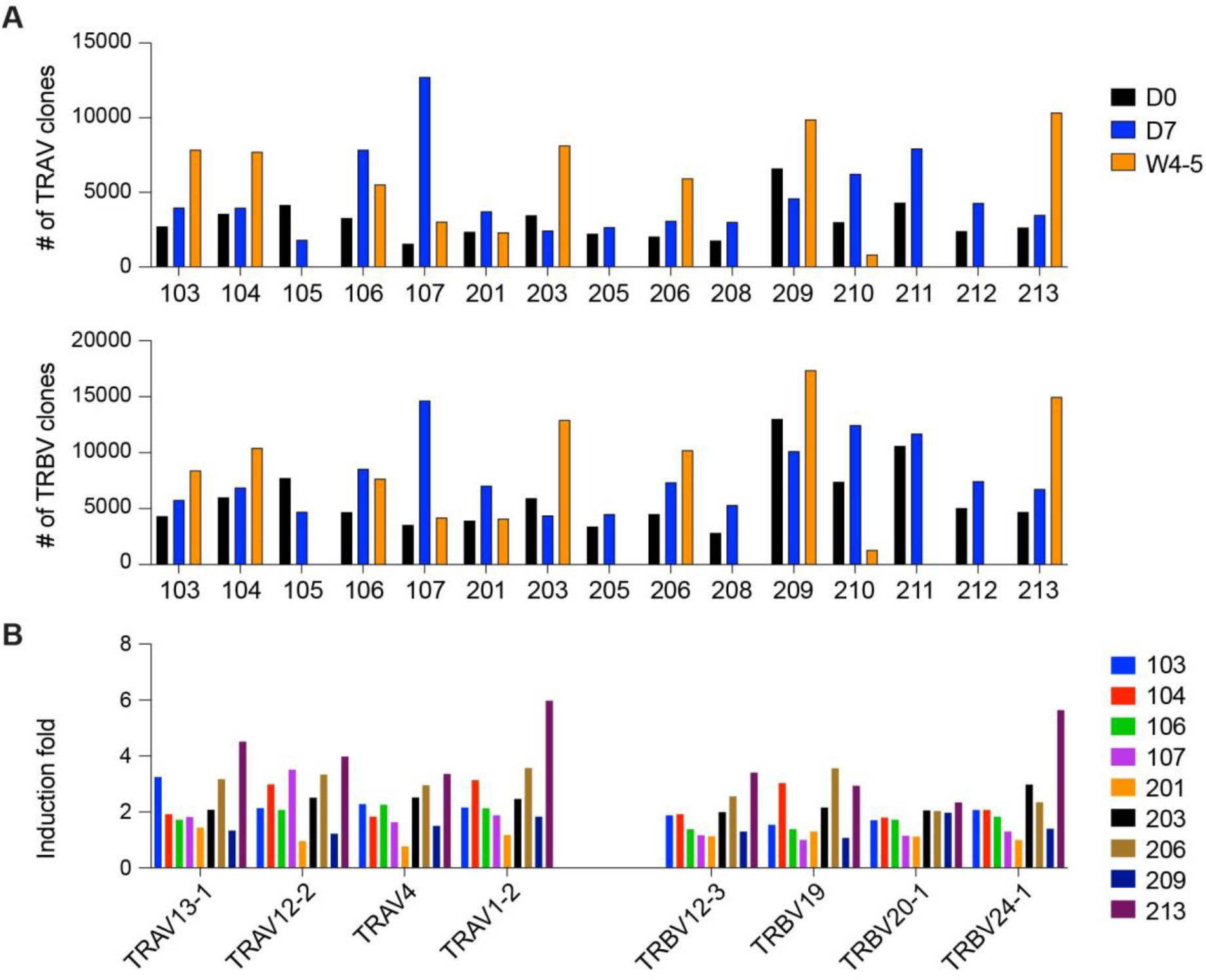
SARS-CoV-2 T-cell memory. (A) Bar graphs were shown the number of clonotypes in each patient after the vaccination. (B) Box plots show the induction fold of variable (V) gene usage from TCRα and TCRβ chain.

## Discussion

In this study, we demonstrate that CLL patients under active surveillance and those that are treatment-naïve exhibit a superior response to COVID-19 vaccination than patients on active treatment. While naïve or minimally treated patients showed an expanded humoral and cellular immune response, the heavily pretreated patients exhibited only T cell immune responses, even with repeated immune stimulation utilizing a heterologous vaccination regimen. The value of a third vaccination was most pronounced in patients who exhibited evidence of a recuperated immune system following effective CLL treatment. These results coincided with pattern of transcriptional expression of immune genes and BCR/TCR repertoire.

Interrogation of the transcriptional response to vaccination utilizing RNA-seq highlighted that nearly all CLL patients demonstrated transcriptional activation of early innate immune response pathways, including interferon-JAK/STAT signaling,^17,25^ within two days, regardless of the antibody response. Interferon-mediated innate immune response serves as a biomarker and plays a critical role in the immune system to control viral replication combating SARS-CoV-2 infection.^17,45,46^Skewed IGHV usage, including the appearance of *IGHV1-69, IGHV 4-34*, and *IGHV 3-21*, was observed in the *BCR* repertoire of the CLL patients. Diverse *IGHV* usage occurs in COVID-19 patients and vaccinated individuals.^17,25^. In addition, the final number of clonotypic B cells detected was much lower in non-IgG responders and even in the seropositive CLL-patients were lower vaccinated healthy controls. The data is consistent with increased numbers of SARS-CoV-2 specific IGHV clones correlating with an improvement of humoral response rate and B-cell reconstitution.

Hypogammaglobulinemia in CLL patients results from leukemic cells perturbating the interaction between T and B cells. Patients with low serum immunoglobulin levels typically show ineffective humoral responses after both primary and subsequent boosting doses.^14,23,47^ Half of patients with persistent immunodeficiency or B-cell depletion following initial vaccination remain seronegative after a booster dose.^8,9,14,48^ T cell immunity is essential for viral recognition and clearance and cellular responses can prevent initial infection and seroconversion.^10,23,48^ T cells are especially critical in immune protection against SARS-CoV-2 in cancer patients, who undergo therapy with B cell depleting agents, such as anti-CD20 antibody.^49,50^ Here we report that CLL patients with diminished numbers of functional CD19+ B cells, a key player in humoral response against SARS-CoV-2 virus, developed robust T cell immune responses to COVID-19 vaccination.

Importantly, we also found a BTKi or BCL2i treatment-dependent effect on the immune response to COVID-19 vaccine boosters. Vaccine effectiveness is moderated by the time between treatment completion and vaccination.^8,51^ Here, CLL patients treated for more than five years failed to seroconvert, while patients pretreated with the BTK inhibitors ibrutinib or acalabrutinib for two years or less seroconverted after 2-3 doses of a COVID-19 vaccine. Prolonged BTKi treatment predisposes patients towards an ineffective vaccine immune response because B-cell maturation relies on functional BTK.^52^ Increased serum IgA levels in BTKi-treated CLL patients appears to improve functional humoral immunity demonstrated by decreased infection susceptibility and hospitalization rates.^2,52^

Protection from severe disease, hospitalization and death by COVID-19 vaccination results from the combination of humoral immunity with a durable cellular immune memory response. In immunocompromised patients the timing of initial and booster vaccination should be carefully considered in reference to the underlying disease status. Remission-inducing therapy resulting in improved immune status and B-cell reconstitution improves adaptive immunity.^12^

## Limitations of the study

There are several limitations to this study. The study was conducted on volunteers from a specific geographical area, Munich (Germany), total numbers of CLL patients were limited, and samples were not available from all patients for all vaccination timepoints for the RNA-seq studies.

## Acknowledgments

Our gratitude goes to the participants who contributed to this study to advance our understanding of COVID-19 vaccination. This work used the computational resources of the NIH HPC Biowulf cluster (http://hpc.nih.gov). RNA-sequencing was conducted in the NIH Intramural Sequencing Center, NISC (https://www.nisc.nih.gov/contact.htm).

This work was supported by the Intramural Research Program (IRP) of the National Institute of Diabetes and Digestive and Kidney Diseases (NIDDK) and the Bavarian State Ministry of Science and Art, Special Program for the Promotion of COVID-19 Research (AKZ H.40001.1.7, to MB).

## Authorship

Contribution: H.K.L., M.A.H., P.A.F., C.M.W. and L.H. designed the research study; H.K.L. performed RNA-seq and data analysis; M.A.H and T.T.P. selected the patients, collected, and curated data; H.K.L, M.A.H., M.B., T.T.P., P.A.F., C.M.W. and L.H. analyzed results; H.K.L. prepared figures; H.K.L. and T.T.P prepared table; M.A.H., M.B., T.T.P., J.W.H., K.M., S.Z., H.V.B., P.G., R.W., L.B. and L.P. prepared samples; H.K.L, M.A.H., M.B., P.A.F., C.M.W and L.H. wrote manuscript. All authors read and approved the final version.

